# Premarital sex and its association with child marriage among ever-married women: a study of national representative survey

**DOI:** 10.1101/2023.02.26.23286461

**Authors:** Md Arif Billah, Md. Mostaured Ali Khan, Syed Manzoor Ahmed Hanifi, M Mofizul Islam, Md. Nuruzzaman Khan

## Abstract

**Background:** Sexual engagement before marriage (premarital sex) can influence child marriage; however, the evidence is scarce in low- and middle-income countries (LMICs), including Bangladesh. This study aimed to explore the association between premarital sex and child marriage among ever-married women aged 15-24 years.

**Methods:** We analysed data of ever-married women aged 15-24 years after extracting from the 2017/18 Bangladesh Demographic Health Survey (BDHS). Premarital sex (yes, no) was the primary exposure in this study, and child marriage was the outcome variable. Multilevel mixed-effect logistic regression models were used to determine the extent to which premarital sex influences the occurrence of child marriage.

**Results:** The prevalence of premarital sex and child marriage was 27% and 76.6%, respectively. Ever-married women who experienced child marriage had higher odds of reporting that they had engaged in premarital sex (aOR: 2.68; 95% CI: 2.20-3.26). The odds of premarital sex for women who experienced child marriage were higher in both urban (aOR: 2.67; 95% CI: 1.88-3.80) and rural areas (aOR: 2.69; 95% CI: 2.11-3.43). Child marriage was more common among women of relatively poor households who had premarital sex than those from other wealth quintiles.

**Conclusion:** Premarital sex significantly impacts girls’ child marriage in Bangladesh, and it varies greatly depending on the household’s socioeconomic status. Expanding existing school and community-based programmes aimed at reducing girls’ child marriage, abstinence from early premarital-sexual intercourse and context-specific multi-component interventions for at-risk young people may reduce these practices.

## Background

Child marriage, marriage before 18 years of age, is a common practice among women in many parts of the world. However, its prevalence is very high least developed countries, where approximately 37% of women aged 20-24 married before turning their ages 18 years and 11% before turning 15 years. This prevalence is roughly twice that in developed countries (UNICEF, 2022a). The rate is further higher in South Asian and African countries (UNICEF, 2022a), even after significant reduction of child marriage occurrence during the Millennium Development Goals period of 2000 and 2015 (UNICEF, 2022a). This relatively high prevalence of child marriage poses a challenge to achieving United Nations Sustainable Development Goal 5.3 for women and girls: abolition of eliminate all harmful practices, such as child, early and forced marriage and female genital mutilations (Lo Forte et al., 2019; UN, 2015).

Child marriage has substantial consequences on the health and development of adolescent girls and their offspring (Elnakib et al., 2022; Kaya et al., 2022; Lichter & McCloskey, 2004; Zabin & Kiragu, 1998). Child brides typically have little awareness of sexual and reproductive health issues. As a result, they may engage in risky sexual behaviour, experience unwanted pregnancies, have high fertility rates and/or experience sexually transmitted infections (STIs) (Fan & Koski, 2022). Many of these adolescent girls become pregnant and give childbirths at ages when they are not physically or mentally prepared and may experience pregnancy complications, poor maternal health, premature births, underweight babies, or even deaths new-borns (Efevbera et al., 2017; Paul et al., 2019). These dangers are heightened twofold since early brides frequently come from marginalized families and have poor nutrition (Chung et al., 2018). Early pregnancy has become the leading cause of death among adolescents and young girls worldwide, with higher incidents in low and middle-income countries (LMICs) (Huda et al., 2021).

Bangladesh has one of the world’s highest prevalence of child marriage (UNICEF, 2021, 2022b), where over 50% of women now in their mid-20s were married before they reached 18 years (UNICEF, 2020). The rate is further higher in low socioeconomic tiers—rural areas, women with little or no formal education and culturally or religiously conservative households (Asadullah et al., 2021; Chowdhury, 2004; Fattah & Camellia, 2022; Henry et al., 2015; Kamal et al., 2015; Saleheen et al., 2021; Scott et al., 2021; Talukder et al., 2020). Child marriage was also found to be more prevalent among Muslim women and women who lived in families with strong patriarchy (Kamal et al., 2015).

Premarital sexual activity occurs prior to formal marriage. Historically, various religions regard this as a moral issue, although it is acceptable in Western countries (Finer, 2007). Premarital sex is culturally forbidden in many settings, and there are strict laws prohibiting it. However, global statistics indicate that 40% and 75% of adolescents between the ages of 15-19 and 20-24 have premarital sex, respectively and its very common in developed countries than LMICs (Tegegne, 2022). For instance, in the USA, 74% women aged 15-44 have had premarital sex by the age of 20 (Finer, 2007). Contrarily, in Sub-Saharan Africa, approximately 54% of girls and 43% of boys had premarital sex before the age of 18 (Melesse et al., 2021). In Bangladesh, One out of ten school going adolescents were engaged in the premarital sexual intercourses in Bangladesh (Murshid & Irish, 2020) and its more prevalence among university students (30%) (2021).

In many LMICs, including Bangladesh, premarital sexual activity is socially and religiously prohibited and a sin. Also, in conservative societies, often premarital sexual intercourse occurs secretly and unplanned manner, increasing the risk of unprotected sexual behaviours and unintended pregnancies (Adhikari et al., 2018; Biratu et al., 2022). Particularly in societies where premarital sex is a religious sin and a social taboo, parents try to marry off their daughters as soon as they become aware of any illicit sexual behaviours. If the girls become pregnant, they are left with no choice but to marry (Horii, 2020) even if they are below 18. Many parents marry off their daughters prior to 18 out of fear of social consequences even if the girls did not have a history of engaging in premarital sex till that point because they want to avert risks (Fagbamigbe & Idemudia, 2017; Glynn et al., 2010). Therefore, it substantially connected to school dropout, particularly among girls as well (Biddlecom et al., 2008). A few evidences show parents’ concern about their daughters engaging in sexual activities before marriage as a potential influencer of child marriage (Lowe et al., 2019; Melesse et al., 2021; Misunas et al., 2021; Stark, 2018; Yakubu & Salisu, 2018). However, very few studies from LMICs have explored the effect of premarital sexual intercourse on child marriage (Misunas et al., 2021; Stark, 2018) using population-level datasets, and none from Bangladesh, to our knowledge. In this study, we explored the effects of premarital sex on the occurrences of child marriage among ever-married adolescent and adult women in Bangladesh. This evidence has high importance in developing interventions to reduce child marriage and protect girls’ sexual and reproductive health rights.

## Methods

### Data source

In this study, we analysed the 2017/18 Bangladesh Demographic Health Survey (BDHS) data. The BDHS is a nationally representative survey conducted every three years as part of the Demographic and Health Survey (DHS) programme of the USA. The National Institute of Population Research and Training (NIPORT), a government organisation under the Ministry of Health and Family Welfare of Bangladesh, conducted this survey. Technical and financial support was provided by several development partners, including UNFPA and UNDP.

### Survey design

The BDHS used a stratified and two-stage cluster sampling design to identify the sample population across the countries. In the first stage, a total of 675 Primary Sampling Units (PSUs) were selected randomly from a list of 293,579 PSUs, generated by the Bangladesh Bureau of Statistics (BSS) as part of the National Population Census 2011. PSU is typically a city block in urban and village(s) in rural areas. Of the 675 selected PSUs, the survey was conducted in 672 and the remaining three PSUs were excluded due to severe floods in those areas. In the second stage, 30 households were selected randomly from each of the selected PSUs by probability proportional to PSU size. This produced a list of 20,160 households, among which interviews were conducted in 19,457. There were 20,370 eligible respondents in the selected households and the conditions of eligibility were: (i) married women aged 15-49 years old and (ii) passed the previous night of the day the survey was conducted in the selected households. With less than a 1% non-response rate, the survey data was finally collected from 20,127 women. Details of the sampling strategy have been published elsewhere (National Institute of Population Research Training, 2020).

### Study sample

The data for this study was extracted from the original sample of the survey based on the following two criteria: (i) ever-married women aged 15-24 years old and (ii) reported their age of marriage and age of first sex. We included women aged only 15-24 years to reduce recall bias and make the estimates relatively recent. A total of 5,596 women met these criteria (Figure 1).

**Figure 1.**
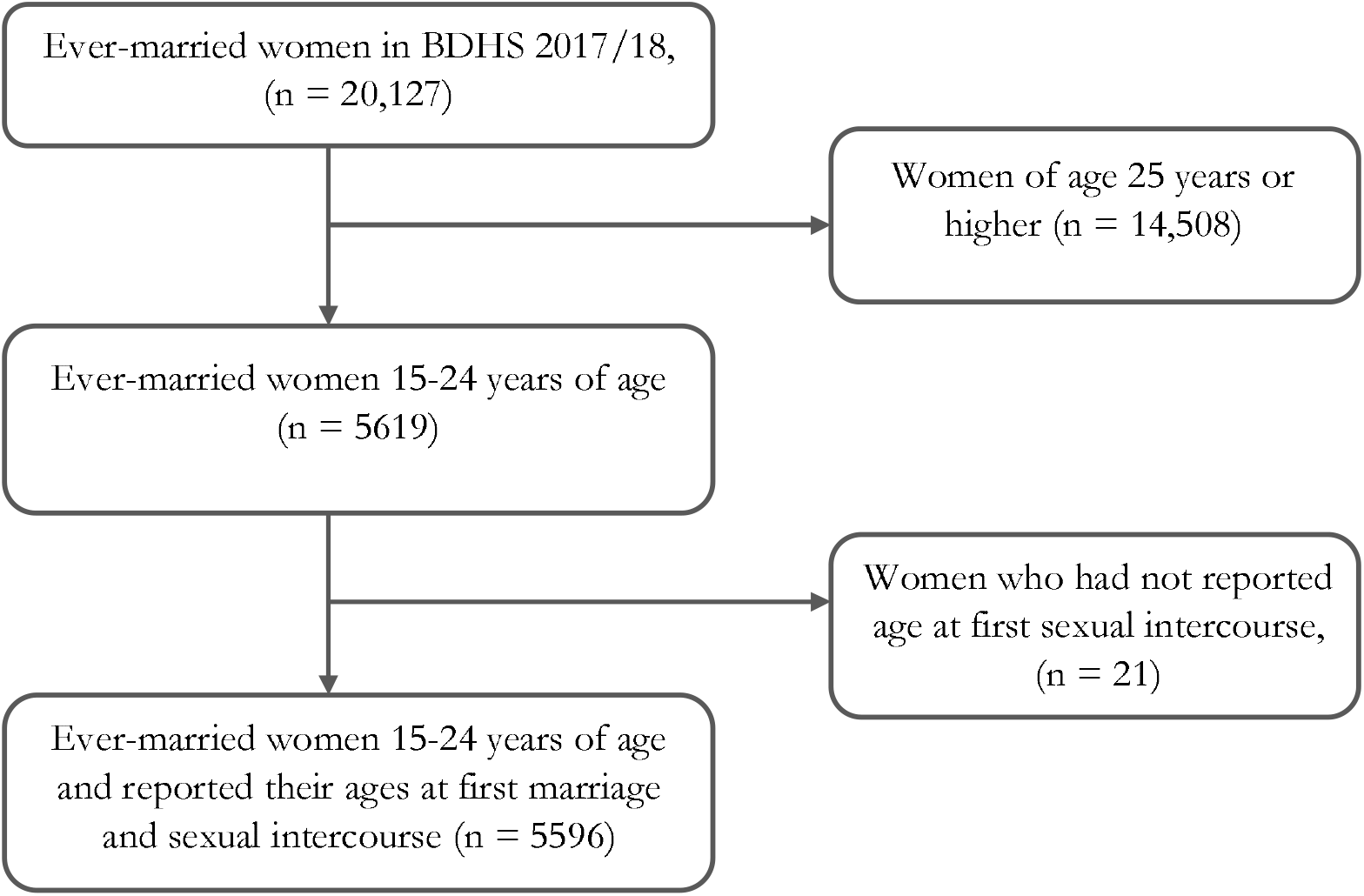
Sample selection process for the study analysis

### Outcomes variable

The outcome variable was child marriage (yes, no). The BDHS recorded this information by asking women, “How old were you when you first started living with him?” We categorised participants’ responses as child marriage (if the marriage occurred before the age of 18) and normal-aged marriage (if the marriage occurred at 18 or later) as per the universal recommendation (UNICEF, 2022a).

### Explanatory variable

The primary explanatory variable was premarital sexual intercourse (yes, no). We developed this binary variable by comparing women’s age at first marriage and age at first sexual intercourse. Due to social norms and religious binding, sex outside marriage is prohibited in Bangladesh. Therefore, women who reported their first sexual intercourse earlier than the age of their first marriage, irrespective of whether it was with their partners who they married or not, were classified as “had premarital sex.” Accordingly, those who experienced first sexual intercourse after their first marriage, irrespective of whether it was with their husbands or not, were identified as “did not have premarital sex”.

### Covariates

We considered a range of covariates to adjust the association between premarital sex and child marriage. We first created a list of relevant variables by reviewing the available literature on child marriage in Bangladesh and neighbouring LMICs and then checked the availability of those variables in the BDHS (Chowdhury, 2004; Fattah & Camellia, 2022; Henry et al., 2015; Hossen & Quddus, 2021; Kamal et al., 2015; Majumdar, 2018; Martin et al., 2001; Melesse et al., 2021; Mokhtari et al., 2022; Murshid & Irish, 2020; Saleheen et al., 2021; Seff et al., 2021; Smith-Greenaway et al., 2021). The listed variables that were available in the BDHS were finally selected as covariates. The selected variables were women’s education (no education, primary, secondary, and higher), religion (Muslim and non-Muslim), household wealth index (poorer, poor, middle, rich and richer), sex of the household’s head (male and female), place of residence (urban and rural) and administrative divisions (Barishal, Chattogram, Dhaka, Khulna, Mymensingh, Rajshahi, Rangpur, and Sylhet). Two community-level variables were included, they were community-level poverty (high poverty, moderate poverty, low poverty and middle/rich community) and community-level illiteracy (low, moderate, and high). These two variables were not directly available in the survey. We calculated them using participants’ individual-level data on wealth quintile and education at the PSU (considered as a community) level. The calculation procedure for these two community-level variables has been published elsewhere (Khan et al., 2020).

### Statistical analysis

Descriptive analysis was used to describe the characteristics of the respondents. Assessment of the relationship between premarital sex and child marriage was carried out by using multilevel mixed-effect logistic regressions. Both adjusted and unadjusted effects of covariates on child marriage were assessed. In Bangladesh, child marriage prevalence varies primarily by residence type and household wealth status (Scott et al., 2021); thus, we conducted subgroup analyses to see the variation in the effect size. For association between premarital sex and child marriage by household wealth status, we ignore community level poverty because it has a higher chance to yield multicollinearity issues. Coefficients were reported with odds ratio (both adjusted and unadjusted) and 95% confidence intervals. Sampling weights were applied in all analyses to account for the complex survey design. The study was designed and reported in accordance with the Strengthening the Reporting of Observational Studies in Epidemiology (STROBE) guidelines (von Elm et al., 2014). STATA version 16 was used for analyses (StataCorp. 2019. Stata Statistical Software: Release 16. College Station, TX: StataCorp LLC).

## Results

### Sample characteristics

The demographic characteristics of 5,596 ever-married women aged 15-24 years are presented in Table 1. Around 63% of them were between the ages of 20 and 24. Approximately 54% of all respondents had completed secondary school and 93% were Muslims. In terms of household wealth, 22.3% belonged to the rich quantile, 20.6% to the middle, and 19.9% to the poor wealth quantile. Over 75% of the sample reported living in rural areas. More than half (57.4%) of the respondents were from communities with moderate literacy levels and nearly half (47.2%) from high poverty levels. Among ever-married women, the prevalence of premarital sex and child marriage was 27% and 76.6%, respectively.

**Table 1.** Characteristics of ever-married women aged 15-24 years in Bangladesh, n = 5596

| Variables | n | % (95% CI) |
| --- | --- | --- |
| **Age groups** |  |  |
| 15-19 | 2053 | 36.7 (35.1-38.3) |
| 20-24 | 3543 | 63.3 (61.7-64.9) |
| **Education** |  |  |
| No education | 176 | 3.2 (2.6-3.8) |
| Primary | 1304 | 23.3 (21.7-25) |
| Secondary | 3051 | 54.5 (52.8-56.2) |
| Higher | 1066 | 19.0 (17.6-20.6) |
| **Religion** |  |  |
| Muslim | 5193 | 92.8 (91.2-94.1) |
| Non-Muslim | 403 | 7.2 (5.7-9.3) |
| **Wealth index of household** |  |  |
| Poorer | 1,023 | 18.3 (16.4-20.3) |
| Poor | 1,116 | 19.9 (18.5-21.5) |
| Middle | 1,153 | 20.6 (19.1-22.2) |
| Rich | 1,249 | 22.3 (20.5-24.3) |
| Richer | 1,055 | 18.8 (17.1-20.7) |
| **Sex of household head** |  |  |
| Male | 4913 | 87.8 (86.7-88.9) |
| Female | 683 | 12.2 (11.1-13.4) |
| **Administrative divisions** |  |  |
| Barishal | 303 | 5.4 (4.9-6.0) |
| Chattogram | 1079 | 19.3 (17.8-20.8) |
| Dhaka | 1497 | 26.8 (25-28.5) |
| Khulna | 586 | 10.5 (9.7-11.4) |
| Mymensingh | 448 | 8.0 (7.3-8.8) |
| Rajshahi | 744 | 13.3 (12.1-14.5) |
| Rangpur | 614 | 11.0 (10.0-12.0) |
| Sylhet | 324 | 5.8 (5.2-6.4) |
| **Place of residence** |  |  |
| Urban | 1536 | 27.5 (26.0-29.0) |
| Rural | 4060 | 72.6 (71.0-74.0) |
| **Community level literacy** |  |  |
| Low (<25%) | 1167 | 20.9 (17.5-24.7) |
| Moderate (25% to 50%) | 3212 | 57.4 (52.8-61.9) |
| High (>50%) | 1217 | 21.8 (18.3-25.7) |
| **Community level poverty** |  |  |
| High (>41%) | 2641 | 47.2 (43.2-51.2) |
| Moderate (16% to 41%) | 1382 | 24.7 (21.0-28.9) |
| Low (<=15%) | 712 | 12.7 (10.1-16.0) |
| Middle/rich | 860 | 15.4 (13.1-18.0) |
| **Premarital sexual intercourse** |  |  |
| No | 4083 | 73.0 (71.5-74.4) |
| Yes | 1513 | 27.0 (25.6-28.6) |
| **Child marriage** |  |  |
| No | 1308 | 23.4 (21.9-24.9) |
| Yes | 4288 | 76.6 (75.1-78.1) |

**Note:** All percentages are weighted. Only ever married women were considered in the analysis, thus, the prevalence of child marriage would vary from the national statistics.

### Distribution of premarital sex and child marriage

Table 2 presents the bivariate distribution of women who had engaged in premarital sexual intercourse and child marriage across respondents’ characteristics. Almost 31% of the respondents who engaged in premarital sexual intercourse were in the 15-19 years age-group and around 32% completed up to secondary education. Approximately 27% of the Muslim respondents, 30.8% of those belonged to richer households and 27.4% from female-headed households had premarital sexual intercourse. Experience of premarital sex was relatively high among the women who lived in Chattogram division (31%) and urban areas (27.3%).

**Table 2.** Bivariate analyses of the characteristics of ever-married women according to premarital sex and child marriage

| Characteristics | Premarital sex, % (95% CI) (n=1,513) | Child marriage, % (95% CI) (n=4,288) |
| --- | --- | --- |
| **Age** |  |  |
| 15-19 years | 30.9 (28.5-33.3) | 90.0 (88.3-91.5) |
| 20-24 years | 24.8 (23.1-26.7) | 68.9 (66.9-70.7) |
| **Education** |  |  |
| No education | 8.9 (5.3-14.4) | 81.5 (74.8-86.8) |
| Primary | 16.2 (14.0-18.8) | 82.8 (80.1-85.3) |
| Secondary | 32.3 (30.3-34.3) | 83.4 (81.6-85.1) |
| Higher | 28.3 (25.2-31.6) | 48.7 (45.3-52.2) |
| **Religion** |  |  |
| Muslim | 27.3 (25.7-28.9) | 77.4 (75.8-78.9) |
| Non-Muslim | 24.2 (19.9-29.1) | 66.7 (61.3-71.6) |
| **Wealth status** |  |  |
| Poorer | 21.6 (19.1-24.4) | 85.9 (83.3-88.1) |
| Poor | 27.3 (24.4-30.5) | 79.6 (76.8-82.0) |
| Middle | 28.3 (25.2-31.6) | 78.5 (75.5-81.3) |
| Rich | 26.9 (24.0-29.9) | 73.3 (70.1-76.3) |
| Richer | 30.8 (27.4-34.4) | 66.3 (63.0-69.5) |
| **Sex of the household head** |  |  |
| Male | 27.0 (25.6-28.5) | 76.3 (74.7-77.8) |
| Female | 27.4 (23.2-32.0) | 78.9 (75.2-82.2) |
| **Place of residence** |  |  |
| Rural | 26.4 (24.0-29.0) | 73.6 (70.8-76.3) |
| Urban | 27.3 (25.5-29.1) | 77.7 (75.9-79.5) |
| **Division** |  |  |
| Barishal | 28.3 (23.7-33.3) | 79.8 (75.5-83.5) |
| Chattogram | 30.9 (27.3-34.8) | 72.8 (69.3-76.1) |
| Dhaka | 24.8 (21.9-28.0) | 75.1 (71.1-78.8) |
| Khulna | 27.9 (23.7-32.6) | 77.5 (73.5-81.1) |
| Mymensingh | 23.2 (19.4-27.5) | 77.3 (73.1-81.0) |
| Rajshahi | 28.8 (24.9-33.1) | 85.9 (82.3-88.8) |
| Rangpur | 27.1 (22.9-31.7) | 82.6 (78.7-85.9) |
| Sylhet | 22.8 (18.9-27.2) | 57.9 (52.7-63.0) |
| **Community level illiteracy** |  |  |
| Low (<25%) | 31.8 (27.9-35.9) | 71.7 (68.5-74.7) |
| Moderate (25% to 50%) | 26.7 (25.0-28.5) | 76.4 (74.3-78.3) |
| High (>50%) | 23.4 (20.5-26.5) | 82.0 (78.7-84.8) |
| **Community level poverty** |  |  |
| High (>41%) | 25.3 (23.3-27.4) | 80.2 (78.0-82.2) |
| Moderate (16-41%) | 28.2 (25.4-31.1) | 78.0 (74.8-80.8) |
| Low (<=15%) | 32.5 (28.2-37.1) | 67.5 (63.0-71.8) |
| Middle/rich | 26.1 (22.4-30.0) | 71.0 (66.7-74.9) |

**Note:** All percentages are weighted and are presented in row percentages. We only considered ever married women in the analysis.

The prevalence of child marriage was 83.4% among women who complete up to secondary education, 77.4% among Muslim women and 85.9% among those from poorer households. Child marriage was higher among women in urban than rural areas (77.7%). The prevalence was relatively high among women who resided in the Rajshahi (85.9%), Rangpur (82.6%) and Barishal (79.8%) divisions. Respondents from the communities with a higher level of illiteracy and poverty reported a higher prevalence of child marriage (82.0% and 80.2%, respectively).

The prevalence of premarital sexual intercourse among women who experienced child marriage was approximately two times higher than it was among those who were married at normal ages, and it holds true across four levels of education (**Figure 2**). Overall, 30.3% of women who experienced child marriage reported having engaged in premarital sexual intercourse compared to 16.5% of women who were married at mature ages. The prevalence of premarital sex was highest among those who received higher education than those who received other lower tiers of education. There appears to have an increasing trend in percentages of women who reported engaging in premarital sex once we move from no formal education to higher education..

**Figure 2.**
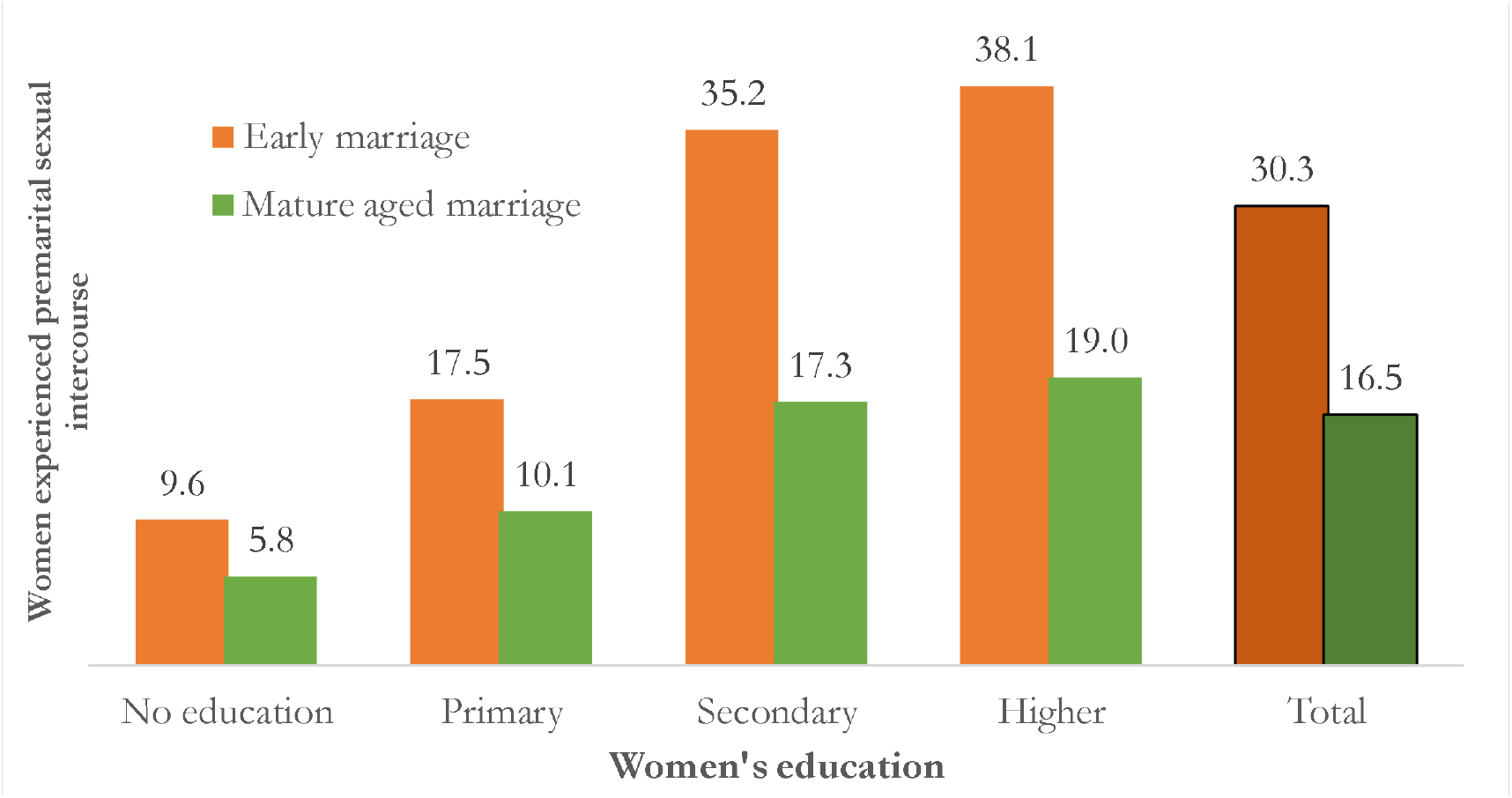
Distribution of premarital sexual experience among early- and normal-aged married women across levels of education, BDHS 2017/18

### Association between premarital sex and child marriage in Bangladesh

Women who experienced child marriage had 2.68 times higher odds of reporting that they had engaged in premarital sex (aOR: 2.68; 95% CI: 2.20-3.26) than women who were married at mature ages (**Table 3**). Sub-group analysis across urban (aOR: 2.67; 95% CI: 1.88-3.80) and rural (aOR: 2.69, 95% CI: 2.11-3.43) areas also produced similar results (**Figure 3**). However, there is a gradual decline in the likelihood of reporting premarital sex among women who experienced child marriage across wealth quintiles except for the richer group. For instance, the odds of engaging in premarital sexual intercourse among the poorer subgroup were 5.37 times (95% CI: 2.33-12.37) among women who were married prior to 18 than those who married at normal ages. The odds ratios decline as we move toward the poor, middle-wealth (aOR: 2.50, 95% CI: 1.55-4.05) and rich subgroups. However, the odds ratio in the richer household subgroup slightly deviates from the trend (aOR, 2.58, 95% CI: 1.79, 3.72). Detailed results of these subgroup analyses are presented in Supplementary Tables 1 and 2.

**Table 3.**
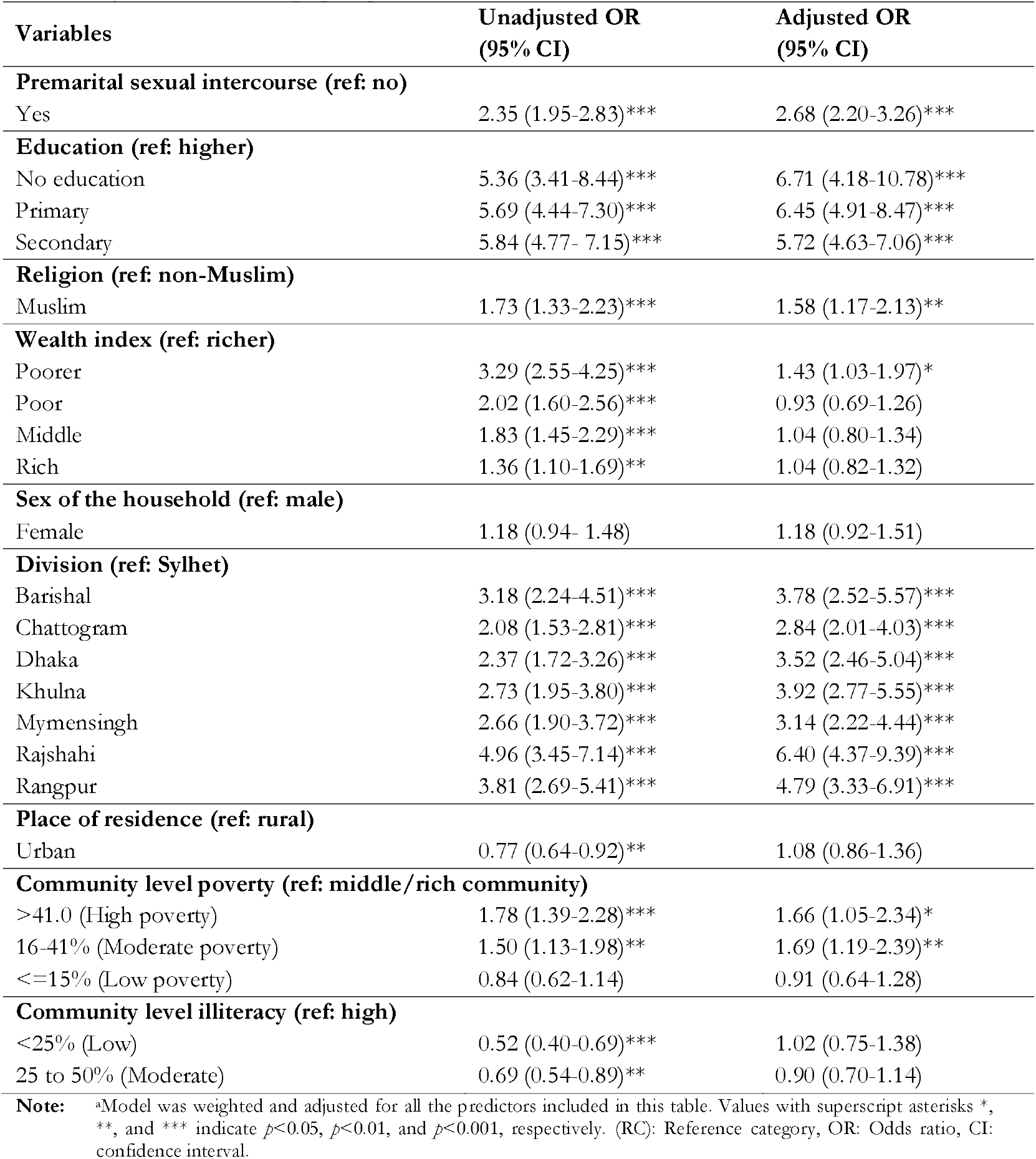
Multi-level mixed-effect bivariate and multivariable logistic regressions examining the associations of premarital sexual intercourse with child marriage, adjusted for the individual- and community-level socio-demographic predictors

| Variables | Unadjusted OR<br>(95% CI) | Adjusted OR<br>(95% CI) |
| --- | --- | --- |
| **Premarital sexual intercourse (ref: no)** |  |  |
| Yes | 2.35 (1.95-2.83)*** | 2.68 (2.20-3.26)*** |
| **Education (ref: higher)** |  |  |
| No education | 5.36 (3.41-8.44)*** | 6.71 (4.18-10.78)*** |
| Primary | 5.69 (4.44-7.30)*** | 6.45 (4.91-8.47)*** |
| Secondary | 5.84 (4.77- 7.15)*** | 5.72 (4.63-7.06)*** |
| **Religion (ref: non-Muslim)** |  |  |
| Muslim | 1.73 (1.33-2.23)*** | 1.58 (1.17-2.13)** |
| **Wealth index (ref: richer)** |  |  |
| Poorer | 3.29 (2.55-4.25)*** | 1.43 (1.03-1.97)* |
| Poor | 2.02 (1.60-2.56)*** | 0.93 (0.69-1.26) |
| Middle | 1.83 (1.45-2.29)*** | 1.04 (0.80-1.34) |
| Rich | 1.36 (1.10-1.69)** | 1.04 (0.82-1.32) |
| **Sex of the household (ref: male)** |  |  |
| Female | 1.18 (0.94- 1.48) | 1.18 (0.92-1.51) |
| **Division (ref: Sylhet)** |  |  |
| Barishal | 3.18 (2.24-4.51)*** | 3.78 (2.52-5.57)*** |
| Chattogram | 2.08 (1.53-2.81)*** | 2.84 (2.01-4.03)*** |
| Dhaka | 2.37 (1.72-3.26)*** | 3.52 (2.46-5.04)*** |
| Khulna | 2.73 (1.95-3.80)*** | 3.92 (2.77-5.55)*** |
| Mymensingh | 2.66 (1.90-3.72)*** | 3.14 (2.22-4.44)*** |
| Rajshahi | 4.96 (3.45-7.14)*** | 6.40 (4.37-9.39)*** |
| Rangpur | 3.81 (2.69-5.41)*** | 4.79 (3.33-6.91)*** |
| **Place of residence (ref: rural)** |  |  |
| Urban | 0.77 (0.64-0.92)** | 1.08 (0.86-1.36) |
| **Community level poverty (ref: middle/rich community)** |  |  |
| >41.0 (High poverty) | 1.78 (1.39-2.28)*** | 1.66 (1.05-2.34)* |
| 16-41% (Moderate poverty) | 1.50 (1.13-1.98)** | 1.69 (1.19-2.39)** |
| <=15% (Low poverty) | 0.84 (0.62-1.14) | 0.91 (0.64-1.28) |
| **Community level illiteracy (ref: high)** |  |  |
| <25% (Low) | 0.52 (0.40-0.69)*** | 1.02 (0.75-1.38) |
| 25 to 50% (Moderate) | 0.69 (0.54-0.89)** | 0.90 (0.70-1.14) |
| **Note:** | aModel was weighted and adjusted for all the predictors included in this table. Values with superscript asterisks *, **, and *** indicate $p<0.05$, $p<0.01$, and $p<0.001$, respectively. (RC): Reference category, OR: Odds ratio, CI: confidence interval. | |

**Figure 3.**
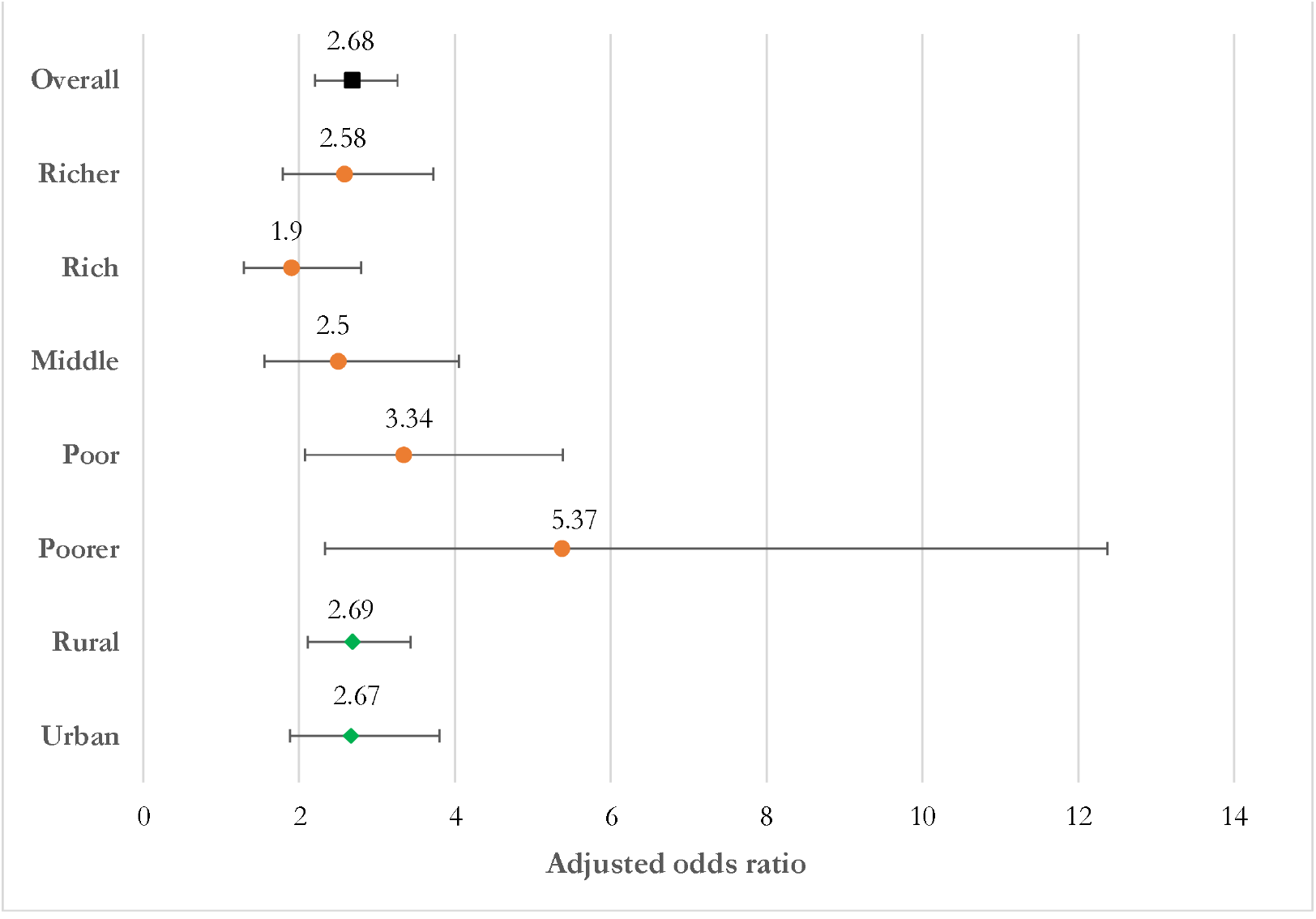
Associations between child marriage and premarital sexual intercourses across wealth quintiles and place of residence

### Other significant factors associated with child marriage in Bangladesh

Several other factors were found significantly associated with child marriage. Women who experienced child marriage had significantly higher odds of receiving no formal education (aOR: 6.71; 95% CI: 4.18-10.78), only primary (aOR: 6.45; 95% CI: 4.91-8.47) or secondary education (aOR: 5.72; 95% CI: 4.63-7.06) than higher education((**Table 3**). Women who experienced child marriage were more likely (aOR: 1.58; 95% CI: 1.17-2.13) to be Muslim than non-Muslim. In terms of wealth quintile, women who experienced child marriage had 1.43 times higher odds that they were from poorer households than richer households. Women in the Rajshahi division had the highest likelihood of child marriage (aOR: 6.40; 95% CI: 4.37-9.39), followed by Rangpur (aOR: 4.79; 95% CI: 3.33-6.91) and Khulna Division (aOR: 3.92; 95% CI 2.77-5.55) compared to women in the Sylhet division. Women who experienced child marriage had higher odds of living in communities of moderate poverty (aOR: 1.69; 95% CI: 1.19-2.39) or high poverty (aOR: 1.66, 95% CI: 1.05-2.34) than in middle or rich communities.

## Discussion

The study aimed to investigate the association between premarital sex and the occurrence of child marriage among Bangladeshi women aged 15 to 24 years. Our findings suggest that three out of every four ever-married women aged 15-24 were married before the age of 18, and one-third of them had sexual intercourse prior to their marriage. Women who experienced child marriage had nearly three times higher odds than those who married at mature ages to report that they had engaged in premarital sexual intercourse. This association was strongest among women from households of comparatively low socio-economic status or living in communities with relatively high poverty. With rapid westernisation in Bangladesh, premarital sexual activities are likely to increase in the coming days, and so are the occurrences of child marriage unless the existing stigma towards premarital sex is drastically reduced.

A quarter of ever-married women reported having premarital sexual intercourse. This estimate is higher than the available estimates for Asian countries (Dave et al., 2013; Mahmoodi et al., 2020) and lower than that for African regions (Behulu et al., 2019; Hailegebreal et al., 2022; Melesse et al., 2021; Seff et al., 2021; White et al., 2000). This finding has a crucial implication, as premarital sexual intercourse is completely prohibited in Bangladesh and many other LMICs. Consequently, women engage in premarital sexual intercourse do so hiddenly mostly using none or less effective contraception methods. Apart from enhancing the occurrences of child marriage, as our findings suggest, premarital sexual intercourse also increases the risk of unintended pregnancies; and subsequent abortion, which is allowed only to save a woman’s life. Unprotected sexual intercourse also increases the risk of catching sexually transmitted infections (Efevbera et al., 2017; Fan & Koski, 2022; Paul et al., 2019; Teferra et al., 2015). Easy access to contraception by all women – married or unmarried - is critical to mitigate these risks. However, it is challenging to implement this in Bangladesh due to social and religious norms.

There could be multiple reasons for the higher odds of child marriage among girls of relatively poor households who had engaged in premarital sex. Though premarital sexual activity is mostly hidden, any indication motivates parents to marry off their daughter because of cultural and religious issues and fear of getting social restriction (Lowe et al., 2019; Melesse et al., 2021; Misunas et al., 2021; Stark, 2018; Yakubu & Salisu, 2018). This practice is even more common among Muslim people (over 90% of the total population in Bangladesh are Muslim) which is supported by this study finding. Poor people are more likely to live in communities with higher levels of no or little formal education. In such a community, premarital sex conveys a relatively high degree of stigma, while child marriage does little or none. On the contrary, wealthy and educated families may take premarital sex relatively lightly and are likely to have the capacity to manage such an adverse situation. Besides, they overlook such engagement by considering their daughter’s age, education and empowerment. It is also found that women engage in premarital sexual activity usually get married within six months of their sexual engagement (Sassler & Lichter, 2020; Sassler et al., 2018).

Poverty driven child marriage is well evident in LMICs and Bangladesh (Ahonsi et al., 2019; Jordana, 2016; Kohno et al., 2020; Melesse et al., 2021; Montazeri et al., 2016; Seth et al., 2018; Tekile et al., 2020). The underlaying reasons are lack of access to girls’ education and reducing economic burden to family related to bearing costs for daughter education and living expenses. Another important reason is security as poor wealth quintile women are mostly resided in the poor community where violence against girls are commonly reported in Bangladesh. Also, individual or community-level poverty may raise the vulnerability of poor households by reducing their abilities to handle the situation arising from daughters’ premarital sexual intercourse. These existing challenges are increase further in occurrence of premarital sex as parents in this case consider their social dignity as much more important and marry off their daughter is an effective solution rather than ignoring risk of child marriage because of their unawareness.

Girls in most LMICs often lack the agency to navigate decision-making around sexual activity, yet society expects them to abstain from sexual activity until they get married and upload their personal and family reputations (Koenig et al., 2020). Child marriage remains the only way to cover up premarital sex and/or pregnancy for parents whose daughters were engaged in premarital sex. Besides, religiosity was seen to support child marriage even if girls did not engage in premarital sex (Mobolaji et al., 2020). Since the government and non-government organisations are increasingly trying the prevent child marriage, some parents then resort to using fake ages for their daughters just to meet the legal age of marriage or delay registering the marriage. Clearly, this is a complex problem. If society continues to hold a stigma on premarital sex and its prevalence increases, strict implementation of the Child Marriage Restraint Act (2017) may have substantial adverse consequences. While self-regulation interventions and counselling may be used to reduce the prevalence of premarital sex, how much these will be effective in this internet era remains unknown.

### Strengths and limitations

This study has several strengths. To our knowledge, this is the first study of its kind that explored the association between premarital sex and child marriage by analysing large and nationally representative data. Advanced statistical techniques were used to explore the associations and adjusted for the individual-, household-, and community-level factors. We also conducted sub-group analyses and explored the associations further. Therefore, the findings reported in this study are precise and can be used for national-level policy and program formulation. However, the major limitation of this study is the use of cross-sectional data, meaning that the results are correlational only, not causal. Furthermore, the data were self-reported, and interviewers were unable to validate the responses. Moreover, since premarital sex is prohibited in Bangladesh, it is possible that many women did not provide accurate information. However, such a bias is likely to be random. Moreover, we only considered ever-married women to explore the relationship between premarital sex on child marriage because the BDHS did not collect data on premarital sex from unmarried women.

## Conclusion

One-third of married women aged 15-24 years had engaged in sexual intercourse prior to their first marriage, and it increased the odds of child marriage. The odds are even higher among women who received no formal education, from households of relatively low wealth quintiles or living in communities of high-level poverty. The findings are likely to be robust and appropriate for informing evidence-based policies and programmes to improve the situation. Existing school and community-based programmes aimed at reducing girls’ child marriage could be expanded further including covering the aspects of early premarital sexual activities. Interventions can be developed involving multi-sectoral stakeholders to promote girls’ formal and moral education, strict enforcement of the legal marriage age, and community awareness that are likely to reduce the relatively high rate of child marriage. However, it is challenging to bring social changes to reduce premarital sexual intercourse, particularly in this internet era.

## Data Availability

Data associated in this study is freely available in https://dhsprogram.com/

## Declarations

## Abbreviations

LMICs: Low-and Middle-Income Countries
DHS: Demographic Health Survey
BDHS: Bangladesh Demographic Health Survey
aOR: adjusted odds Ratio
CI: Confidence Interval
SDG: Sustainable Development Goal
NIPORT: National Institute of Population Research and Training
PSU: Primary Sampling Unit

## Authors’ contribution

MAB, MMAK and MNK developed the study concepts and analysed the data. MAB and MMAK drafted the manuscript. MNK, MMI and SMAH critically reviewed the manuscript. All authors have seen and approved the final version of the paper.

## Competing interest

None.

## Funding

The study has no external or institutional fund.

## Data availability

Data associated in this study is freely available in https://dhsprogram.com/

## Ethics approval and consent to participate

The data of this study were obtained from MEASURE DHS Archive, and originally collected by the Macro, Calverton, USA. ORC Macro Institutional Review Board reviewed and approved the data collection procedure. Informed consent was obtained from each participant prior to enrolment.

## Availability of data and materials

The dataset can download after registering with the MEASURE DHS at: http://dhsprogram.com/data/Using-DataSets-for-Analysis.cfm.

## Acknowledgement

We are thankful to MEASURE DHS for the data support and also grateful to icddr,b is also grateful to the Governments of Bangladesh, Canada, Sweden and the UK for providing core/unrestricted support for its operations and research, where the data for this study was analysed. The authors also acknowledge the support of Health System and Population Studies Division of icddr,b and Department of Population Science of Jatiya Kabi Kazi Nazrul Islam University, where this study was designed and conducted.

## Supplementary files

**Supplementary Table 1.**
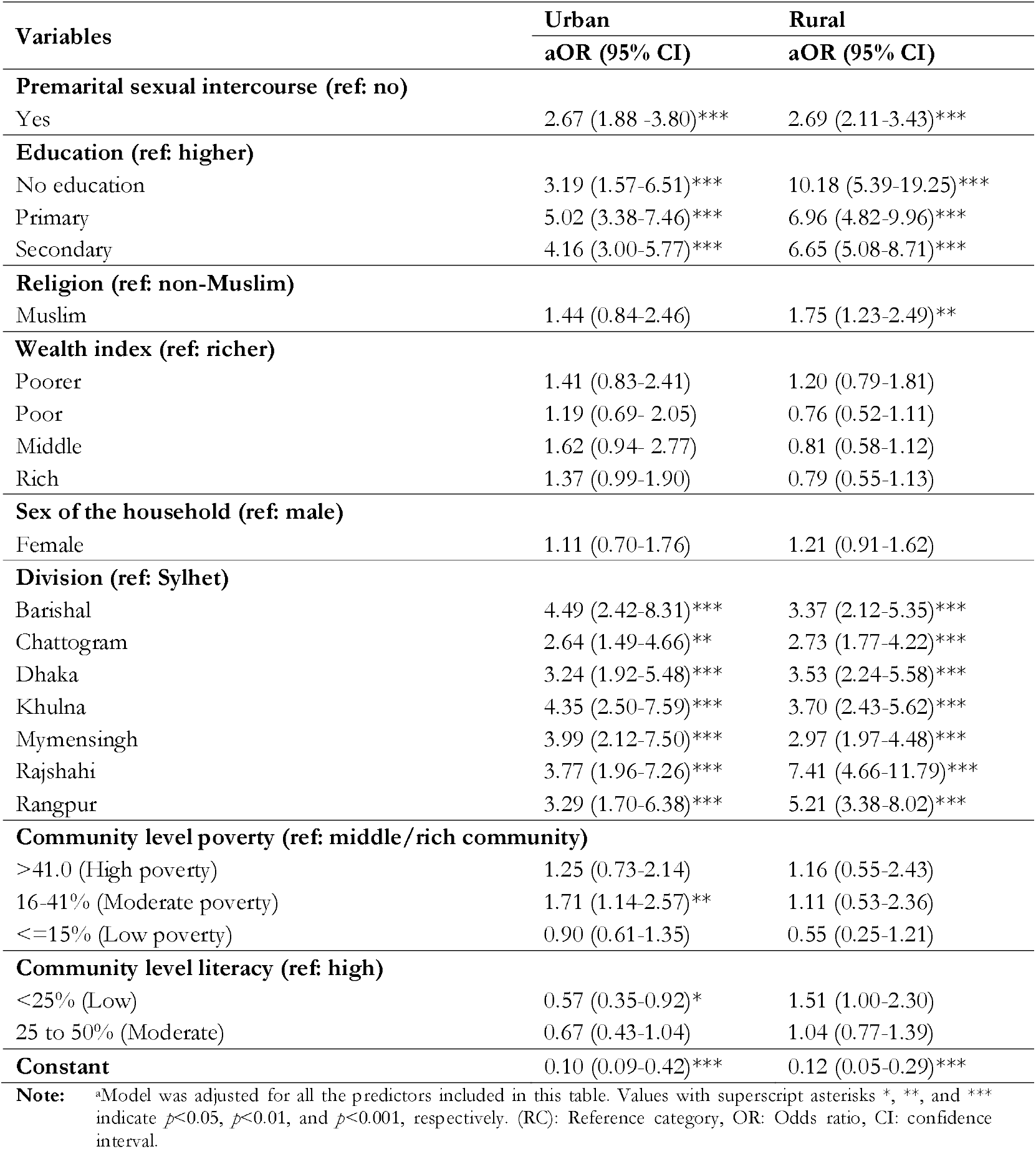
Urban-rural difference in association of premarital sex with child marriage adjusted with individual and community level socio-demographic predictors using multi-level mixed effect multivariate logistic regression.

**Supplementary Table 2.** Household wealth status difference in association of premarital sex with child marriage adjusted with individual and community level socio-demographic predictors using multi-level mixed effect multivariate logistic regression†.

| Variables | Poorer<br>aOR (95% CI) | Poor<br>aOR (95% CI) | Middle<br>aOR (95% CI) | Rich<br>aOR (95% CI) | Richer<br>aOR (95% CI) |
| --- | --- | --- | --- | --- | --- |
| **Premarital sexual intercourse (ref: no)** |  |  |  |  |  |
| Yes | 5.37 (2.33-12.37)*** | 3.34 (2.07-5.38)*** | 2.50 (1.55-4.05)*** | 1.90 (1.29-2.79)** | 2.58 (1.79-3.72)*** |
| **Education (ref: higher)** |  |  |  |  |  |
| No education | 11.52 (4.36-30.45)*** | 7.20 (2.60-19.93)*** | 6.15 (1.64-23.10)** | 2.26 (0.78-6.54) | 19.58 (2.16-177.65)** |
| Primary | 8.71 (3.88-19.57)*** | 6.07 (3.14-11.71)*** | 7.61 (4.09-14.18)*** | 4.67 (2.79-7.82)*** | 5.85 (3.09-11.07)*** |
| Secondary | 8.04 (3.70-17.47)*** | 5.39 (3.18-9.13)*** | 5.66 (3.55-9.034)*** | 4.78 (3.23-7.09)*** | 5.64 (3.80-8.38)*** |
| **Religion (ref: non-Muslim)** |  |  |  |  |  |
| Muslim | 1.78 (0.92-3.45) | 1.71 (1.01-2.90)* | 1.55 (0.77-3.10) | 1.84 (0.98-3.42) | 1.05 (0.57-1.93) |
| **Place of residence (ref: rural)** |  |  |  |  |  |
| Urban | 0.98 (0.56-1.73) | 0.89 (0.56-1.44) | 0.83 (0.49-1.40) | 0.83 (0.57-1.21) | 1.56 (1.13-2.16)** |
| **Sex of the household (ref: male)** |  |  |  |  |  |
| Female | 0.89 (0.46-1.71) | 0.87 (0.50-1.52) | 0.90 (0.51-1.59) | 1.93 (1.15-3.22)* | 1.52 (0.96-2.41) |
| **Division (ref: Sylhet)** |  |  |  |  |  |
| Barishal | 3.01 (1.63-5.55)*** | 3.18 (1.57-6.43)** | 5.37 (2.24-12.90)*** | 3.18 (1.28-7.90)* | 4.36 (1.76-10.82)** |
| Chattogram | 1.90 (0.90-4.02) | 1.67 (0.85-3.29) | 2.43 (1.17-5.04)* | 2.47 (1.26-4.84)** | 4.73 (2.56-8.74)*** |
| Dhaka | 4.88 (1.79-13.34)** | 1.43 (0.74-2.76) | 4.35 (1.97-9.62)*** | 3.17 (1.64-6.12)** | 4.43 (2.44-8.04)*** |
| Khulna | 2.35 (0.99-5.55) | 5.12 (2.38-10.99)*** | 5.08 (2.31-11.16)*** | 2.76 (1.41-5.37)** | 4.29 (2.12-8.68)*** |
| Mymensingh | 4.71 (2.32-9.57)*** | 1.91 (1.07-3.39)* | 2.98 (1.40-6.35)** | 3.17 (1.46-6.90)** | 4.70 (1.98-11.19)*** |
| Rajshahi | 8.61 (3.64-20.36)*** | 3.62 (1.89-6.92)*** | 8.31 (3.83-18.03)*** | 6.59 (3.05-14.21)*** | 6.74 (3.20-14.17)*** |
| Rangpur | 8.89 (4.52-17.51)*** | 2.90 (1.66-5.05)*** | 5.15 (2.28-11.66)*** | 2.92 (1.27-6.73)* | 4.03 (1.55-10.48)** |
| **Community level illiteracy (ref: high)** |  |  |  |  |  |
| <25% (Low) | 1.52 (0.71-3.24) | 0.73 (0.40-1.34) | 1.80 (0.97-3.34) | 0.57 (0.34-0.97)* | 0.65 (0.36-1.15) |
| 25 to 50% (Moderate) | 0.89 (0.56-1.41) | 1.09 (0.73-1.63) | 1.04 (0.65-1.68) | 0.71 (0.46-1.11) | 0.69 (0.40-1.19) |
| **Constant** | 0.12 (0.03-0.41)** | 0.21 (0.08-0.56)** | 0.14 (0.04-0.42)** | 0.24 (0.09-0.61)** | 0.14 (0.05-0.38)*** |

**Note:** \*Model was adjusted for all the predictors included in this table. Values with superscript asterisks \*, \*\*, and \*\*\* indicate $p < 0.05$ , $p < 0.01$ , and $p < 0.001$ , respectively. (RC): Reference category, OR: Odds ratio, CI: confidence interval. †For association between premarital sex and child marriage by household wealth status, we ignore community level poverty because it has a higher chance to yield multicollinearity issues

## Notes

### Competing Interest Statement

The authors have declared no competing interest.

